# Potential COVID-19 vaccination opportunities in primary care practices in the United States

**DOI:** 10.1101/2021.09.20.21263757

**Authors:** Sanjay Basu, Rebecca Weintraub, Ishani Ganguli, Russell Phillips, Robert Phillips, Asaf Bitton

## Abstract

Rapid, widespread COVID-19 vaccination is critical to pandemic mitigation and recovery. To help policymakers interested in further enhancing primary care delivery of COVID-19 vaccines, it is important to estimate the absolute number of vaccination opportunities, and identify how these opportunities may fall disproportionately among different communities given the unequal way that COVID-19 falls upon communities of color, low-income, and rural communities. To quantify the potential benefits of greater primary care engagement in vaccination efforts, we estimated the number of potential vaccination opportunities (PVOs) in primary care in the remaining calendar months of year 2021, and the possible uptake if we supplied enough vaccine to primary care practices to fulfill their opportunities. To estimate how many potential vaccination opportunities (PVOs) may occur in primary care, we used three sets of data, analyzing the latest available waves of the following: (i) the National Ambulatory Medical Care Survey (NAMCS, 2016, N = 677 providers); (ii) the National Health Interview Survey (NHIS, 2018, N = 29,839 individuals in 29,839 households); and (iii) the Medical Expenditure Panel Survey (MEPS, 2018, N = 40,025 individuals in 14,500 households). Per the NAMCS data, which provide a nationally-representative sample of ambulatory care visits, primary care physicians normally provide 40.2 million primary care visits per month. The majority of the primary care utilization is absorbed by those aged 16 to 64 years old who are not otherwise priority groups (i.e., not having chronic diseases as defined by ACIP) but the second large group of visits are those with a chronic disease (27.2% of all visits). As compared to the NAMCS data providing an estimate of care from the perspective of providers, the overall sample in NHIS provides a view of primary care access and utilization from a population perspective. Per NHIS, 34% of the civilian US population saw a generalist physician in the prior calendar year, or 109.8 million people. Overall, we would estimate that over the latter half of calendar year 2021, approximately 15 million potential vaccine opportunities per month would be available through US primary care practices.

## Introduction

Rapid, widespread COVID-19 vaccination is critical to pandemic mitigation and recovery. To date, efforts to distribute COVID-19 vaccines have been federally- or state-run, with centralized administration to hospitals, mass vaccination sites, pharmacies, and community health centers (e.g., Federally Qualified Health Centers), with limited supply distributed to individual primary care clinics, such that most primary care practitioners were not providing COVID-19 vaccines as of the time of this writing in July 2021.^1^ As vaccination through mass sites abates, and vaccination moves into a “last mile delivery” phase to reach herd immunity by delivering the vaccine to zones with limited access to date, reaching pediatric and adolescent populations, or convincing unvaccinated adults to receive the vaccine, primary care delivery of the COVID-19 vaccines in of increasing interest. Primary care-based vaccine delivery has several potential benefits, including ability to leverage trusting relationships and well-tested vaccination distribution logistics pipeline. Additionally, primary care visit rates have increased and largely rebounded from their nadir during the “shelter-in-place” period of the COVID-19 pandemic, with pent-up demand likely to result in excess utilization of primary care services over the next several months.^2^

Prior studies of vaccination opportunities via primary care practices have estimated the share of non-COVID vaccinations that have been delivered during routine office visits. In the first quarter of 2021, the Robert Graham Center released a white paper analysis of previous immunization delivery patterns, by cataloguing delivery of vaccinations by provider type using 2017 Medicare Part B Fee-For-Service data and the 2013-2017 Medical Expenditure Panel Survey. They noted: “In 2017 Medicare Part B Fee-For-Service, Primary Care Physicians provided the largest share of services for vaccinations (46%), followed closely by Mass Immunizers (45%), then NP/PAs (5%). The Medical Expenditure Panel Survey showed that Primary Care Physicians provided most clinical visits for vaccination (54% of all visits).”^3^

To help policymakers interested in further enhancing primary care delivery of COVID-19 vaccines, it is important to estimate the absolute number of vaccination opportunities, and identify how these opportunities may fall disproportionately among different communities given the unequal way that COVID-19 falls upon communities of color, low-income, and rural communities. To quantify the potential benefits of greater primary care engagement in vaccination efforts, we estimated the number of potential vaccination opportunities (PVOs) in primary care in the remaining calendar months of year 2021, and the possible uptake if we supplied enough vaccine to primary care practices to fulfill their opportunities.

## Methods

To estimate how many potential vaccination opportunities (PVOs) may occur in primary care, we used three sets of data, analyzing the latest available waves of the following: (i) the National Ambulatory Medical Care Survey (NAMCS, 2016, N = 677 providers); (ii) the National Health Interview Survey (NHIS, 2018, N = 29,839 individuals in 29,839 households); and (iii) the Medical Expenditure Panel Survey (MEPS, 2018, N = 40,025 individuals in 14,500 households).

First, the NAMCS data allowed us to estimate the total number of visits to primary care providers during usual demand periods. The NAMCS data were also subset by race/ethnic subgroup and by the common groupings identified in Advisory Committee on Immunization Practices (ACIP) guidelines (e.g., people aged 16-64 with chronic diseases). Additionally, the NAMCS data were subgrouped to compare the typical utilization rates for all types of visits versus vaccination visits among primary care providers and other providers by clinic type and specialty. For the purposes of the analysis, primary care was defined using the National Center for Health Statistics definition as general practice, family practice, general internal medicine or pediatrics.^4^

Second, the NHIS data were used to identify how often people in the community saw a primary care provider. While the NAMCS data are collected from the perspective of the provider and their visits, the NHIS data allowed us to look at the whole civilian US population, and define the probability of overall primary care utilization across the population and its various subgroups, which helps contextualize the NAMCS data in terms of how the utilization seen in primary care corresponds to coverage of the overall civilian population among different population subgroups.^5^ In particular, NHIS is thought to capture minority populations more comprehensively as compared to NAMCS. However, NHIS defines primary care utilization by asking individuals whether they visited a generalist physician during the past year. As a result, NHIS may underestimate primary care utilization because non-physicians (e.g., nurse practitioners) are not included, and as many individuals do not define their primary care physician as a generalist (e.g., some individuals do not define their internal medicine or pediatrics provider as being a generalist^6^).

Finally, the MEPS data were used to understand the distribution of PVOs by insurance type, which is not as reliably recorded in NAMCS or NHIS as in MEPS. MEPS defines a “usual place for care” in terms of emergency room, or non-emergency outpatient clinics either within hospitals or at free-standing facilities, and therefore does not differentiate precisely between primary versus specialty care. Nevertheless, the MEPS data supplement NAMCS and NHIS by providing a sense of how PVOs vary by insurance type.^5,7^

Overall, the three sources of data complement each other to enable us to understand typical primary care reach in terms of the supply of PVOs provided by practices, the population-level reach of primary care, and the disparities implications by race/ethnic subgroup and by insurance type.

All survey sample analyses were weighted to produce nationally-representative estimates. Less than 8% of any single variable was missing, and missing data were not imputed.

## Results

### Usual primary care volume

Per the NAMCS data, which provide a nationally-representative sample of ambulatory care visits, primary care physicians normally provide 40.2 million primary care visits per month. Typically each unique individual patient visits the practice 1.2 times in the survey months (fall and winter, which tends to have higher visit rates), and at the time of this writing (on July 7, 2021) 55.1% of people in the United States of all ages had received at least one dose of the vaccine^8^), leaving 15.0 million PVOs per month.

Figure 1 provides the breakdown of visits by race/ethnic subgroup, while also referencing the population size of that subgroup in the US population as a whole,^8,9^ and providing details of these visits in terms of whether they were visits for vaccinations only or for primary care more generally. Notably, a disproportionate number of Hispanics utilize vaccination visits and utilize primary care visits generally as compared to their population size. By contrast, non-Hispanic Blacks utilize more vaccination visits as compared to their population size, while non-Hispanic Whites utilize fewer vaccination visits but more general primary care than would be proportionate to their population size.

**Figure 1.**
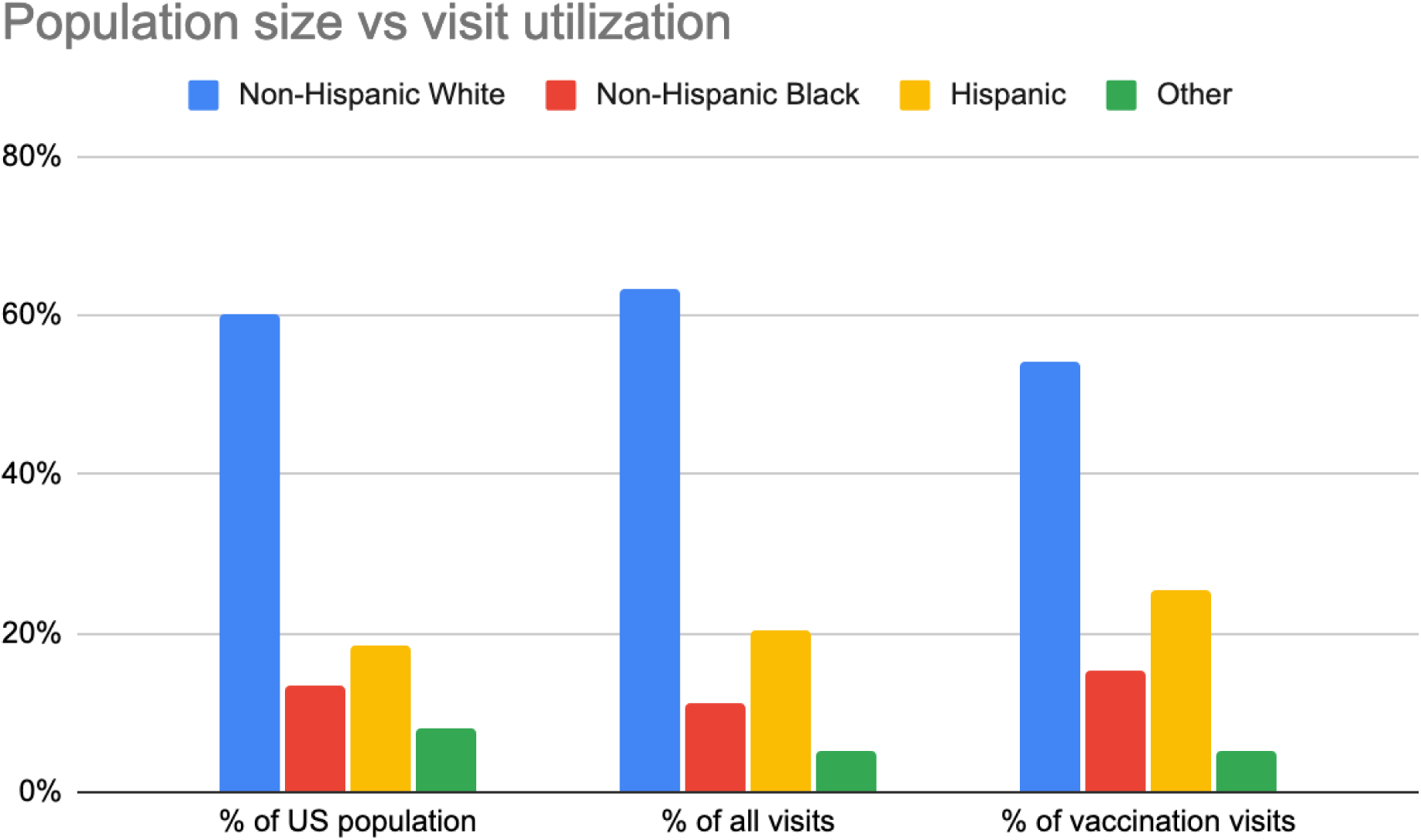
Population size versus visit utilization, based on weighted sample estimates from the National Ambulatory Medical Care Survey (NAMCS, 2016) and US Census population estimates (2016).

Table 1 provides further details of how the groups of particular interest per ACIP guidelines typically appear in primary care. Notably, the majority of the primary care utilization is absorbed by those aged 16 to 64 years old who are not otherwise priority groups (i.e., not having chronic diseases as defined by ACIP) but the second large group of visits are those with a chronic disease (27.2% of all visits). Those over age 65 years and smokers aged 16-64 years old make up the next largest share of visits.

**Table 1.** Proportion of key populations of interest to the Advisory Committee on Immunization Practices for COVID-19 vaccination, based on weighted sample estimates from the National Ambulatory Medical Care Survey (NAMCS, 2016).

|  | Proportion of population age 16+ years | Visits/yr to primary care | Visits with a vaccine administered | Proportion of visits with vaccine administered |
| --- | --- | --- | --- | --- |
| Age 65+ | 27.0% | 88,569,839 | 2,084,842 | 2.4% |
| Age 18-64 with chronic disease* | 23.3% | 131,092,347 | 2,189,739 | 1.7% |
| Age 16-64 with cancer | 0.3% | 507,835 | 0 | 0.0% |
| Age 16-49 and pregnant | 1.3% | 10,490,245 | 0 | 0.0% |
| Age 16-64 and smoker | 20.0% | 88,471,716 | 1,727,305 | 2.0% |
| Others age 16-64 | 28.1% | 162,831,243 | 17,406,739 | 10.7% |

### Population coverage

As compared to the NAMCS data providing an estimate of care from the perspective of providers, the overall sample in NHIS provides a view of primary care access and utilization from a population perspective. Per NHIS, 34% of the civilian US population saw a generalist physician in the prior calendar year, or 109.8 million people.

Figure 2 shows the overall fraction of different race/ethnic groups that reported having seen a generalist physician in the past year, and compares the distribution to the latest estimates (as of June 30, 2021) of COVID-19 vaccination (at least one dose) by race/ethnic group.^10^ Notably, more non-Hispanic Black individuals than non-Hispanic White or Hispanic individuals reported seeing a generalist physician in NHIS; in the vaccination data, by contrast, non-Hispanic Blacks were the least vaccinated among other race/ethnic groups.

**Figure 2.**
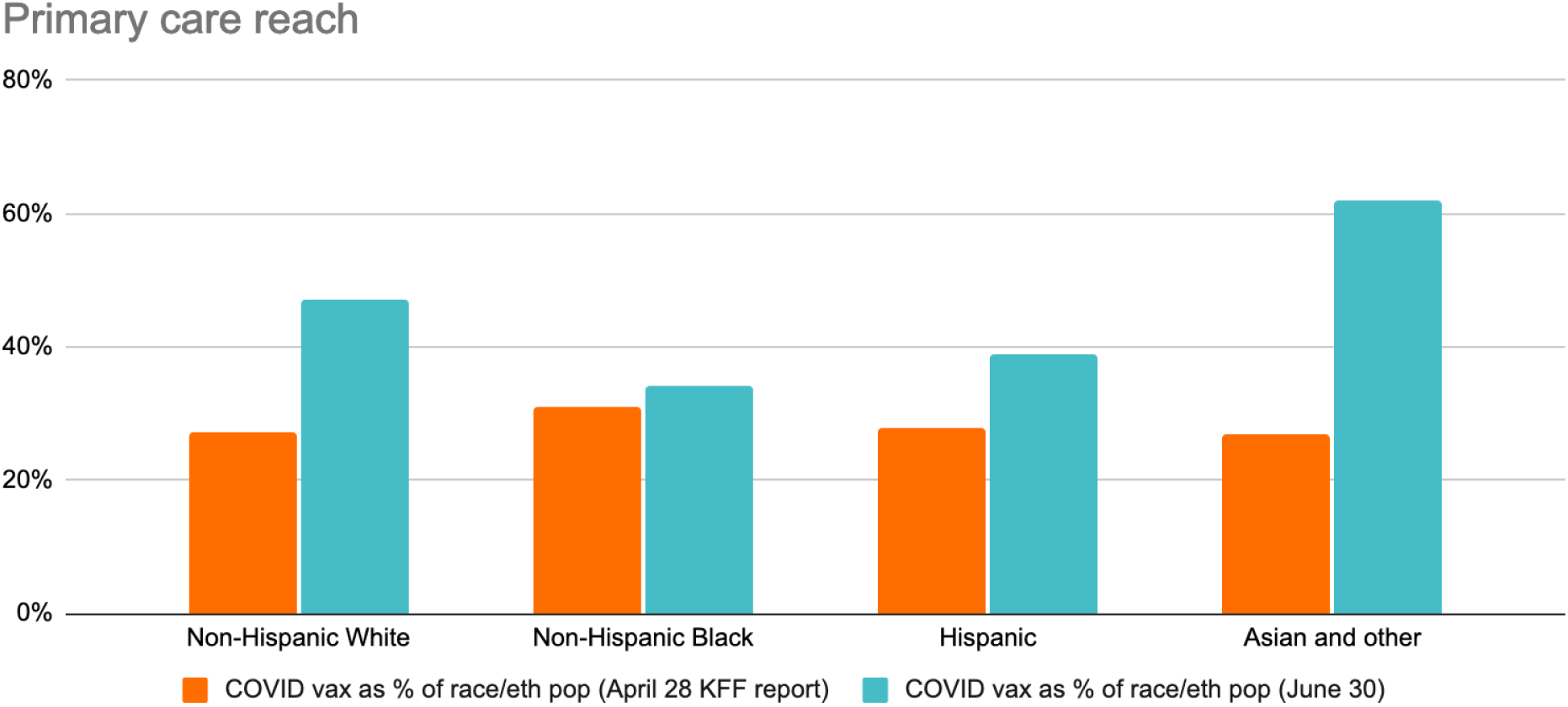
Proportion of people who saw a generalist physician in the National Health Interview Survey (2018), by race/ethnic group, versus estimated rates of COVID-19 vaccination as of June 30, 2021.

### Utilization by insurance type

Table 2 summarizes the proportion of people in MEPS, by age, who had a usual place for care, and--of those--the subsets who had a non-hospital clinical versus a hospital clinic or emergency room as their usual place for care.

**Table 2.** Weighted estimates of populations with a usual place for care, and the proportion of those with a usual place for care who receive it through an emergency room, hospital outpatient clinic, or non-hospital clinic, per the Medical Expenditure Panel Survey (2018).

| Age: | < 65 years old |  |  | 65+ years old |  |  |  |  |
| --- | --- | --- | --- | --- | --- | --- | --- | --- |
| Insurance: | Any private | Public only | Uninsured | Medicare only | Medicare and Private | Medicare and Other Public | Uninsured | No Medicare but any Public/Private |
| Have a usual place for care? | 73.9% | 78.0% | 38.2% | 88.5% | 91.9% | 86.4% | 15.7% | 85.7% |
| Usual place is: |  |  |  |  |  |  |  |  |
| Emergency room | 0.2% | 0.7% | 2.5% | 0.5% | 0.3% | 0.1% | 23.8% | 0.0% |
| Hospital outpatient clinic | 31.7% | 36.8% | 30.2% | 31.8% | 28.5% | 38.0% | 48.2% | 40.0% |
| Non-hospital clinic | 58.6% | 56.2% | 59.1% | 53.2% | 58.6% | 49.5% | 0.0% | 57.0% |

Notably, among those less than 65 years old, over one-third of the uninsured had a usual place for care, and the majority of those were in non-hospital clinics. Additionally, nearly 80% of those less than 65 years old and on public insurance had a usual place for care, with the majority again through non-hospital clinics.

Among those over 65 years old, around 90% of those receiving any type of Medicare coverage had a usual place for care, with about half of those through non-hospital clinics.

## Discussion

We would estimate that over the latter half of calendar year 2021, approximately 15 million PVOs would be available through US primary care practices. Notably, we would expect that these PVOs would be disproportionately utilized by non-Hispanic Blacks and Hispanic Americans, which may address the disproportionate impact of COVID-19 on minority communities.^11^ Given systemic racism, its implications for mistrust of healthcare institutions, and its association with COVID-19 vaccine receipt,^12^ the high trust placed in primary care providers as compared to other healthcare personnel may be particularly important in reaching targeted vaccination rates.^13^

As of April 2021, 89% of primary care providers responding to an Internet survey reported they are ready and willing for their practice to be a vaccination site, but 78% reported not being included as a potential site by their health department, local hospital or health system.^13,14^ The barriers to ensuring entry of primary care providers into the vaccine distribution pool should be further addressed, and include infrastructure and equipment limitations that may be mitigated through the availability of newer cold storage packages and the recent establishment of longer shelf-life for available vaccines,^15–17^ electronic registration and reporting infrastructure that may be more streamlined and accessible to providers,^18,19^ and--perhaps most of all--the inclusion of primary care representatives on state and local vaccine distribution committees and coordination agencies.^20^ The international experience of vaccine distribution has highlighted that several other nations, particularly those in the UK, Europe, and East Asia, have made use of distribution through primary care practices as a backbone for accelerating vaccination towards herd immunity levels.^21^

There are important limitations to the data and conclusions we derive here. First, the datasets are each limited in different ways, but all are limited to the civilian US population, and therefore do not account for important institutionalized populations who may be at particularly high risk for COVID-19 infection. Dedicated surveys of vaccination capacity, distribution, and gaps in care are ongoing for incarcerated and nursing home populations who have experienced particularly high risk.^22,23^ Second, the data are dated and historical, and therefore do not capture the recent shift towards utilization of telehealth visits, which has declined in recent months since the peak of the pandemic, but has converged to a steady-state level that remains higher than prior to COVID-19.^2^ The utilization of telehealth may lower vaccination opportunities in primary care practices, but conversely may permit conversations to be had between practitioners and patients around COVID-19 vaccination before scheduling an in-person vaccination follow-up visit.

Despite these limitations, our findings indicate the potential for last-mile delivery of COVID-19 vaccinations through primary care practices, particularly for minority populations, and suggest that further engagement of primary care practices would be potentially important for national, state, and local COVID-19 vaccination coordinators.

## Data Availability

All data are publicly accessible at.

https://www.meps.ahrq.gov/mepsweb/

https://www.cdc.gov/nchs/ahcd/ahcd_products.htm

https://www.cdc.gov/nchs/nhis/index.htm

